# Subset scanning for multi-trait analysis using GWAS summary statistics

**DOI:** 10.1101/2023.07.19.23292708

**Authors:** Rui Cao, Evan Olawsky, Edward McFowland, Erin Marcotte, Logan Spector, Tianzhong Yang

**Affiliations:** Division of Biostatistics, University of Minnesota, 420 Delaware St. SE, 55455, MN, USA; Technology and Operations Management, Harvard Business School, Soldiers Field, 02163, MA, USA; Division of Epidemiology and Clinical Research, Department of Pediatrics, University of Minnesota, 420 Delaware St. SE, 55454, MN, USA

**Keywords:** Multi-trait analysis, Genome-wide association study (GWAS), Summary statistics, Biobank, Childhood cancer

## Abstract

Multi-trait analysis has been shown to have greater statistical power than single-trait analysis. Most of the existing multi-trait analysis methods only work with a limited number of traits and usually prioritize high statistical power over identifying relevant traits, which heavily rely on domain knowledge. To handle diseases and traits with obscure etiology, we developed TraitScan, a powerful and fast algorithm that agnostically searches and tests a subset of traits from a moderate or large number of traits (e.g., dozens to thousands) based on either individual-level or summary-level genetic data. We evaluated TraitScan using extensive simulations and found that it outperformed existing methods in terms of both testing power and trait selection when sparsity was low or modest. We then applied it to search for traits associated with Ewing Sarcoma, a rare bone tumor with peak onset in adolescence, among 706 traits in UK Biobank. Our analysis revealed a few promising traits worthy of further investigation, highlighting the use of TraitScan for more effective multi-trait analysis as biobanks emerge. Our algorithm is implemented in an R package ‘TraitScan’ available at https://github.com/RuiCao34/TraitScan.

## 1 Introduction

Genome-wide association studies (GWAS) have successfully improved the understanding of the genetic basis of many traits. The emergence of deeply phenotyped GWAS databases such as UK Biobank (Bycroft et al., 2018), eMERGE (Gottesman et al., 2013), and Vanderbilt BioVU (Roden et al., 2008), has facilitated studying associations between single nucleotide polymorphisms (SNPs) and a large number of traits. Phenome-wide association studies (PheWAS) have utilized this rich source of SNP-trait relationships to explore disease risks (Denny et al., 2010) and drug development (Diogo et al., 2018). By evaluating each trait individually, PheWAS is computationally fast to implement. However, it is well documented that joint multivariate analyses can be more powerful than univariate analyses such as PheWAS (Robinson et al., 2018). Many efforts have been devoted to multi-trait analyses that evaluate the relationships between a SNP and a set of traits simultaneously (O’Reilly et al., 2012; Kim et al., 2015; Zhu et al., 2015; Li and Zhu, 2017). Nonetheless, existing multi-trait analyses rely on domain knowledge to select a small number of related traits, and most of them only focus on obtaining high statistical power in hypothesis testing.

We are motivated to understand which risk factors contribute to childhood cancers, which are universally rare and have obscure disease etiology. For example, the etiology of Ewing sarcoma (EWS) remains unclear (Lahat et al., 2008), and conventional epidemiological methods to understand the disease are limited due to the extremely rare incidence rate of EWS (Spector et al., 2021). Recently, several genetic risk factors were identified in a GWAS of EWS (Machiela et al., 2018), potentiating the use of PheWAS to explore the risk factors agnostically. However, PheWAS can suffer from limited statistical power when scanning over a large number of traits, and simply applying the existing multi-trait methods to a large number of traits does not always yield meaningful results as the polygenic nature of some traits would eventually drive the statistical significance. To address the aforementioned limitations, we propose a novel multi-trait analysis method, TraitScan, that values both trait selection performance and high statistical power. Our method ‘TraitScan’ is based on a fast subset scan framework (Neill, 2012) with a linear scan time of the number of traits and thus can handle high-dimensional trait selection. Our method contains three test statistics: higher criticism (HC), truncated chisquare (TC), and a combined test of HC and TC. We note a similar method ASSET (Bhattacharjee et al., 2012) with the same objective. However, it requires an exhaustive search of all possible subsets, which results in an exponential scan time and thus is not computationally efficient when the number of traits exceeds a few dozen.

Our proposed TraitScan algorithm is able to utilize summary-level GWAS data in situations where individual-level data are not available. Due to the logistical limitations and privacy concerns of sharing individual-level data, it has become a common practice to share GWAS summary statistics. Leveraging publicly available summary-level data, our method can filter relevant pleiotropic traits on any given SNP. Through simulations, we show that our method has high power and sensitivity in terms of trait selection under moderately sparse to sparse situations (i.e., the number of truly associated traits is smaller than or close to the square root of total traits). We evaluate traits associated with EWS through GWAS summary statistics on 706 traits filtered from the UK Biobank study (Sudlow et al., 2015). Besides single SNPs, we also show that our method can be extended to genetic scores, such as the predicted gene expression levels in transcriptome-wide association studies (TWAS) (Feng et al., 2021) and polygenic risk scores (PRS) (Torkamani et al., 2018). We implement our method in an R package ‘TraitScan’ that is publicly available at https://github.com/RuiCao34/TraitScan. The package provides an option to use the pre-calculated null distributions of the test statistics, which can handle screening 706 traits in 36 seconds.

## 2. Materials and methods

### 2.1 Models

Our method performs a scan for traits under the null hypothesis that none of the traits is associated with the genetic variant of interest (i.e., SNP). Assume that the data consist of *p* continuous traits and a minor allele dose for a SNP collected from *n* individuals, for *j* = 1, …, *p*. Let *y*_*j*_ = (*y*_1*j*_, …, *y*_*nj*_)^*T*^ be a vector of values of the *j*^*th*^ trait for each of the *n* individuals, *x* = (*x*_1_, …, *x*_*n*_)^*T*^ a vector of the minor allele doses of the SNP of interest, and *ϵ*_*j*_ = (*ϵ*_1*j*_, …, *ϵ*_*nj*_)^*T*^ a vector of error terms for the *j*^*th*^ trait. For continuous traits, we assume that each trait can be modeled as a linear function of the genetic variant, and without loss of generality, *y*_*j*_ is centered and standardized

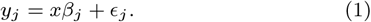

We define our null hypothesis as:

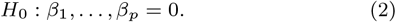

Let **Y** = (*y*_1_, …, *y*_*p*_), ***β*** = (*β*_1_, …, *β*_*p*_) and ***ϵ*** = (*ϵ*_1_, …, *ϵ*_*p*_). We can stack the models together as:

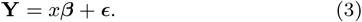

To model the correlation structure between the traits, we assume that *x* is fixed and the rows of ***ϵ*** represent *n × p* i.i.d. observations from an *MV N* (**0, Ω**) distribution, where **0** is a vector of zeroes of length *n × p* and **Ω** = *I*_*n×n*_ ⊗ **Σ**, where **Σ** is the potentially unknown *p × p* covariance matrix of the traits. When the individual-level data are available, the matrix can be estimated from the residuals of fitting separate linear regression models: 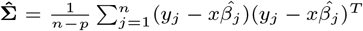, where 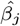 is the ordinary least square estimate.

When only summary statistics are available, which typically consist of 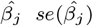 from Equation 1 and the z-score from a Wald test 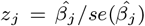 each entry of 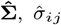 can be approximated using the null SNPs (i.e. the SNPs with no association with any traits) by ignoring the estimation error of 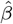 (Kim et al., 2015; Liu and Lin, 2018):

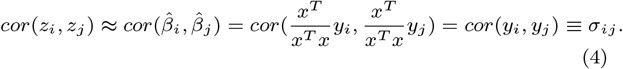

Additionally, when the summary statistics come from overlapping but not identical samples, the correlation between z-scores is still proportional to the trait correlation *σ*_*ij*_ (Li et al., 2021):

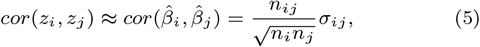

where *n*_*i*_, *n*_*j*_, and *n*_*ij*_ are the sample sizes of trait *i*, trait *j*, and their overlapping samples respectively. Since the (*i, j*)^*th*^ z-score correlation is a constant across all null SNPs, it can be estimated empirically (Zhu et al., 2015):

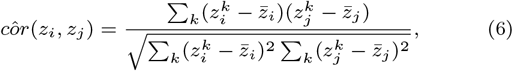

where 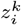 is the z-score for null SNP *k* and trait *i*, and 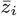 is the mean of vector 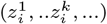 across all null SNPs.

For binary traits, a logistic regression model is usually fitted in GWAS:

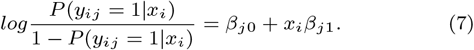

When the effect size *β*_*j*1_ is small, which most often happens in GWAS, the logistic regression model can be approximated by a linear regression model based on the first-order Taylor expansion on *β*_*j*1_:

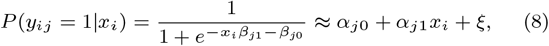

where *α*_*j*0_, *α*_*j*1_, and *ξ* can be regarded as linear regression coefficients. The covariance for binary traits can be similarly derived using summary statistics of the null SNPs. In practice, null SNPs can be chosen based on GWAS p-values (i.e., *>* 0.05).

### 2.2 Scan Statistics

Our subset scanning algorithm relies on a score statistic *F*, which is a function of a non-empty subset *S* ⊆ {1, …, *p*}. It quantifies the amount of anomalousness found in traits {*y*_*j*_ |*j* ∈ *S*} under the null hypothesis that no trait is associated with the SNP. The most anomalous subset is found by maximizing *F* (*S*) over all non-empty subsets of the traits. Calculating *F* (*S*) over all possible subsets *S* is extremely burdensome when *p* is large. To ensure efficient maximization, we use subset scanning techniques and strive for a statistic accompanying priority function that satisfies the strong linear time subset scanning (LTSS) property:

**Definition 1** (Neill (2012)) The score function *F* (*S*) and priority function 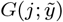 satisfy the strong LTSS property if and only if, for all *j* = 1, …, *p*, max 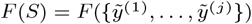, where 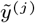 is the trait with the *j*th highest value of 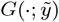.

If *F* (*S*) satisfies the strong LTSS property, the subset *S*^*∗*^ that maximizes *F* (*S*) must be the subset containing the *c* highest-priority traits 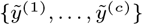 for some *c* between 1 and *p*. Thus, to solve the global optimization problem, we can simply sort the traits by their priority value given by *G* and then compute *F* (*S*) with *S* taken to be one of the *p* subsets 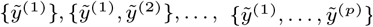.

Neill (2012) gave a constructive theorem that produces a specific priority function 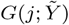 that follows directly from the score function *F* (*S*) when certain properties hold. This pair of functions is then guaranteed to satisfy the strong LTSS property.

**Theorem 1** (Neill (2012)) *Let F* (*S*) = *F* (*T*, |*S*|) *be a function of one additive statistic of subset* 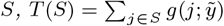 *(where* 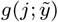 *depends only on trait* 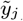 *) and the cardinality of S*, |*S*|. *Assume that F* (*S*) *is monotonically increasing with T* (*S*), *then F* (*S*) *satisfies the strong LTSS property with priority function* 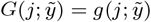.

We thus construct two score statistics, HC and TC that satisfy the conditions of Theorem 1 while quantifying the amount of anomalousness found in a subset of the traits under the null hypothesis. We show by simulations that the HC or TC method had better performance than performing PheWAS under different scenarios. We also combine the two tests by taking the minimum p-value of the two tests, which enables us to achieve results comparable to the better-performed HC or TC method in terms of statistical power and trait selectivity.

#### 2.2.1 Decorrelation

As in Theorem 1, a trait-level statistic is required to quantify the amount of association between a trait and a genetic variant. We used the p-value *p*_*j*_ of the Wald test *z*_*j*_ from the separate regression models (Equation 1), for *j* = 1, …, *p*. As the traits are correlated and sampled from the same or overlapping individuals in our framework, the *p*_*j*_ ‘s are correlated. Herein, we perform the ZCA-cor whitening method (Kessy et al., 2018) on *z*_*j*_, ensuring that the whitened 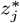 remains maximally correlated with *z*_*j*_. Then we obtain the 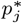 corresponding to 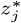.

#### 2.2.2 Higher Criticism Statistic

Following McFowland et al. (2013), we choose the HC score function: 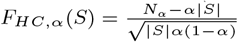, where 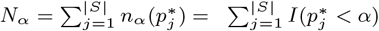). The score function is the standardized difference between the observed count of p-values lower than a p-value threshold *α* and the expected count. According to Theorem 1, it can be easily seen that *F*_*HC*_ (*S*) is a function of |*S*| and one additive statistic *N*_*α*_. *F*_*HC*_ (*S*) is monotonically increasing with *N*_*α*_ and thus satisfies the strong LTSS property with priority function 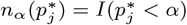.

As we do not know the optimal *α*, we define grid-based HC teststatistic *H*_*HC*_ :

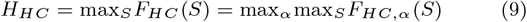

over a grid of *α* and its corresponding subset

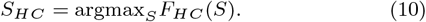

An ideal *α* grid should ensure all possible subsets in the search space, therefore, there should be no more than one p-value between two arbitrary adjacent *α*’s. In practice, we recommend the *α* grid as a geometric sequence from the Bonferroni significant p-value threshold to overall Type I error with a sequence length of 200, i.e. 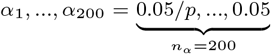. The lower bound of *α* ensures that our test would always be more powerful than PheWAS in special scenarios where all traits are uncorrelated. In the meanwhile, we use an upper bound of 0.05 to decrease the search space.

#### 2.2.3 Truncated Chi-squared Statistic

The HC test may not have ideal performance under non-sparse scenarios (Barnett et al., 2017) and does not take into account the strength of association. To make our method more robust, we propose an additional statistic, which is similar to the truncated z-score method (Bu et al., 2020) and also meets the strong LTSS property. First, we define *γ* as the |*z*^*∗*^| threshold, which is closely related to *α* in HC statistics: *γ* = Φ^*−*1^(1 *− α/*2) and Φ as the cumulative distribution function of a standard normal distribution. The score function *F* (*S*) is defined as *F*_*T C*_ (*S*) =*−log*(*P*_*Mγ*_ (*S*)|*H*_0_), i.e. the negative log p-value of the subset score function 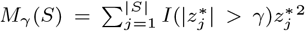 with priority function 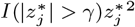.

Note that *M*_*γ*_ is a non-decreasing function of *S*, and *F*_*T C*_ (*S*) is monotonically increasing with *M*_*γ*_, thus satisfying the strong LTSS property. Similarly, we test a grid of *γ* one-to-one mapping to the grid of *α* defined previously:

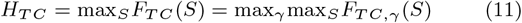

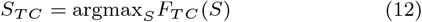

*γ* grid is chosen in correspondence with the *α* values.

### 2.3 Assessing Significance

We have now obtained two subsets *S*_*HC*_ and *S*_*TC*_ that maximize HC or TC statistics. As described above, both subsets always include at least one trait. To determine whether the selected subset is sufficiently anomalous, we calculate the corresponding p-values *p*_*HC*_ and *p*_*T C*_ by comparing the two statistics *H*_*HC*_ and *H*_*T C*_ with their distributions under the null hypothesis.

*p*_*HC*_ can be calculated analytically (Supplementary Materials). As for *p*_*T C*_, we use Monte Carlo (MC) simulations. We start by simulating *p* z-scores under the null, i.e. standard normal distribution for *B* iterations. For the *b*^*th*^ iteration, the *H*_*T C,b*_ can be calculated, and empirical p-values *p*_*T C*_ can be estimated from the simulated *H*_*T C*_ distribution.

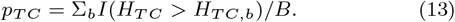

To combine the HC and TC tests, we compare the p-values *p*_*HC*_ and *p*_*T C*_ and get the grid-based statistics:

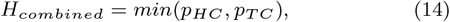

and the traits selected by the combined test are also determined by the test with a smaller p-value:

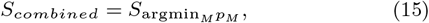

where M = HC or TC. The empirical null distribution of *H*_*combined*_ can be similarly simulated by MC, and the p-value from the combined test *p*_*combined*_ is calculated by comparing the test statistic *H*_*combined*_ and its distribution under the null.

If the null hypothesis is rejected, we can conclude that the SNP is associated with at least one trait contained in *S*^*∗*^. Note that this MC simulation step only depends on the number of traits *p* and the choice of F-statistics. For SNPs sharing the same number of traits *p*, we do not need to recompute the test statistic distribution under the null.

### 2.4 Extension to Genetic Scores

Genetic scores integrate information from multiple SNPs. Linking traits with genetic scores could bring in more statistical power and provide a meaningful interpretation of the results. Genetic scores, which are usually the linear combinations of allele counts of multiple SNPs, have been extensively developed and distributed. Polygenic risk scores that predict the risk of clinical and epidemiological traits (Lewis and Vassos, 2020) or imputation models for gene expression levels in TWAS (Xu et al., 2023) are two types of commonly used genetic scores. We will show how TraitScan can be easily utilized on genetic scores using summary-level GWAS data and an external genetic reference panel.

#### 2.4.1 Continuous Traits

Let *X*_*gs*_ denote the genetic score from *q* SNPs: 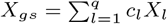, where *X*_*l*_ = (*x*_1*l*_, …, *x*_*nl*_) is the genotype vector for *n* individuals at the *l*^*th*^ SNP, and *c*_*l*_ is the SNP weight vector. In GWAS models, the *j*^*th*^ trait *y*_*j*_ is marginally regressed on each SNP *X*_*l*_, and regression coefficients 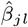 and *se* 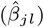 are estimated from the linear regression model

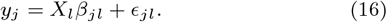

For the genetic score, we are interested in the regression model

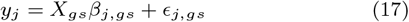

and test the null hypothesis

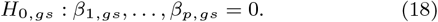

When individual-level data are available, the regression coefficients 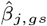 and *se*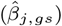 with z statistic from the Wald test 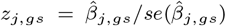 can be directly calculated. When only summary-level data are available, we have

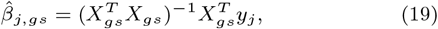

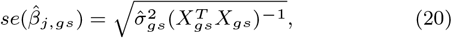

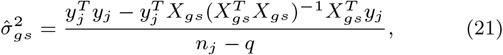

where 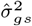 is the residual variance estimate and *n*_*j*_ is the sample size for *j*^*th*^ trait. The items 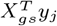 and 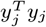 can be derived from GWAS summary data (Pattee and Pan, 2020):

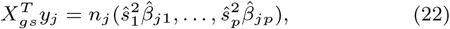

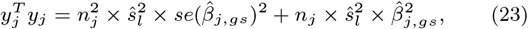

where 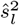 is the variance of SNP *l*. Both 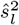 and the genotype matrix 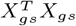 can be estimated from a reference panel comprising genotypic data of individuals from a general population (1000 Genomes Project Consortium, 2015). In practice, for Equation 23, we can calculate *y*^*T*^ *y*_*j*_ across multiple SNPs and take the median as the estimate.

After z statistics *{z*_*j,gs*_*}* are computed, we could follow the same decorrelation and trait scanning steps as above since the genetic score can also be treated as a SNP, and the covariances between *{z*_*j,gs*_*}* are identical under the null hypothesis.

#### 2.4.2 Binary Traits

We have the logistic regression for binary traits:

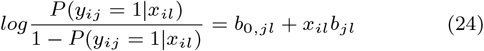

for *i*^*th*^ individual, *j*^*th*^ trait, and *l*^*th*^ SNP, and *b*_0,*jl*_ and *b*_*jl*_ are the regression coefficients. Following Pattee and Pan (2020), we could approximate *P* (*y*_*ij*_ = 1|*x*_*il*_) as a continuous outcome under a linear regression model and denote *β*_0,*jl*_ and *β*_*jl*_ as the coefficients.

The following equations hold:

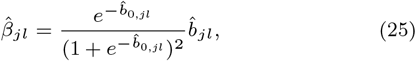

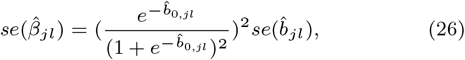

where 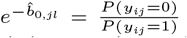 is the ratio of control and case sizes. The logistic regression coefficients can be thus converted to linear regression coefficients and handled by the steps mentioned above.

## 3 Real Data application

We used our method on UK Biobank GWAS data to find out potential traits linked to EWS. EWS is a type of rare childhood cancer in bone or soft tissue (Li and Chen, 2022). Previous studies (Postel-Vinay et al., 2012; Machiela et al., 2018) suggested that six SNPs rs113663169, rs7742053, rs10822056, rs2412476, rs6047482, and rs6106336 were significantly associated with EWS in individuals of European ancestry. We analyzed these six SNPs using the GWAS summary statistics of the UK Biobank data (team, 2020).

UK Biobank is a large-scale database encompassing a broad range of phenotypes, where individuals’ genetic data are linked to electronic health records and survey measures (Sudlow et al., 2015). The phenotypes include population characteristics, biological markers, medical history, environments, dining habits, cognitive functions, etc. In the GWAS study, samples with sex discordance and SNPs with low minor allele counts or low imputation scores were filtered out. Summary statistics of GWAS were obtained from fitting generalized mixed models with a kinship matrix as a random effect and covariates as fixed effects within each genetic ancestry. The heritability of each trait was provided, which was estimated by the Scalable and Accurate Implementation of GEneralized mixed model (SAIGE) (Zhou et al., 2018). More method and analysis details can be found on the Pan-UK Biobank website (https://pan.ukbb.broadinstitute.org/). In our analysis, we focused on the GWAS summary statistics for individuals of European ancestry and with a sufficient number of participants of both genders. We applied the following criteria which left us with 706 traits to perform the TraitScan algorithm:

- Continuous traits with a sample size of at least 5,000 or binary traits with a sample size of at least 5,000 cases and 5,000 controls.
- Traits with genetic heritability estimated to be larger than 0.
- Traits with at least one genome-wide significant SNP (p-value*<* 5 *×* 10^*−*8^).
- Traits with the sample size of each sex larger than 20.
- Traits belonging to these categories were included: health-related outcomes, online follow-up, biological samples, X-ray absorptiometry (DXA), cognitive function, verbal interview, touchscreen questions (except traits related to eyes), and baseline characteristics.

As we intended to evaluate six SNPs, the significance level for TraitScan tests was set at 0.05*/*6 = 0.0083 after the Bonferroni correction. SNPs rs113663169, rs10822056, rs2412476, rs6047482, and rs6106336 were shown to have significant associations with at least one trait out of 706 examined traits in UKBiobank by TraitScan (TraitScan combined tests p-value *≤* 1 *×*10^*−*4^). For SNP rs7742053, TraitScan combined test p-value was 0.863 and thus did not reach statistical significance. Table 1 summarizes the results of TraitScan combined test for the most significant trait in each category identified for the five SNPs. It showed traits that were highly significant in PheWAS were also captured by TraitScan. In fact, TraitScan identified a total of 21 UK Biobank traits related to the five EWS-linked SNPs, while 8 of the trait-SNP associations did not reach statistical significance in PheWAS (using a Bonferroni-significant threshold at 0.05*/*(706 *×* 6) = 1.18*×*10^*−*5^). A full list of selected traits is shown in Supplementary Materials (Table S1).

**Table 1.** Use TraitScan to search among 706 traits in UK Biobank for EWS-linked SNPs.

| SNP | Trait category | Most significant trait in the category | PheWAS p-value |
| --- | --- | --- | --- |
| rs113663169 | Touchscreen questions | Natural hair color: blonde | $\leq 10^{-20}$ |
| | Baseline characteristics | Seated height | $3.19 \times 10^{-7}$ |
| rs10822056 | Biological samples | Monocyte count | $1.19 \times 10^{-13}$ |
| rs2412476 | Biological samples | Aspartate aminotransferase | $\leq 10^{-20}$ |
| | | Erythrocyte distribution width | $\leq 10^{-20}$ |
| rs6047482 | Biological samples | Urea | $1.02 \times 10^{-7}$ |
| rs6106336 | Biological samples | Insulin-like growth factor 1 | $6.16 \times 10^{-7}$ |

To demonstrate the TraitScan application on genetics scores, we further carried out our method on the transcriptomic scores of gene *KIZ* and gene *RREB1*, i.e., the SNP-imputed gene expression of the two genes, with the 706 UK Biobank traits. *KIZ* and *RREB1* had strong evidence to be linked to three top genome-wide significant SNPs in EWS GWAS (*KIZ* was linked to SNPs rs6047482 and rs6106336, and *RREB1* to rs7742053) (Machiela et al., 2018). The weights in the transcriptomic scores of human blood were obtained from Xu et al. (2023), and the internal *r*^2^ of the scores were 0.206 and 0.031 for *KIZ* and *RREB1*, respectively. The significance levels for both genes were set at 0.05*/*2 = 0.025 after the Bonferroni correction. In addition, we also tested the relationship between the number of risk alleles identified in Machiela et al. (2018) with EWS. It was shown that EWS cases had on average 1.08 more risk alleles than controls (p-value = 2.44 *×* 10^*−*63^).

After applying TraitScan on the genetic scores, neither of the two genes *KIZ* (p-value = 0.0826) and *RREB1* (p-value= 1) reached statistical significance in TraitScan, although the trait insulin-like growth factor 1 (IGF-1) picked up by *KIZ* was marginally significant. On the other hand, imputed gene expression of *RREB1*, of which rs7742053 was an expression quantitative trait loci (eQTL), had no evidence of linking to any of the 706 traits. The genetic score of six EWS SNPs, however, was significantly associated with three traits: blonde hair color, ease of skin tanning, and monocyte percentage (TraitScan p-value *<* 1 *×* 10^*−*4^), while the PheWAS identified two additional traits on the EWS score: patient care technician location and facial pain experienced in last month.

To investigate the causal relationship between EWS and the selected traits, we further performed bidirectional Mendelian randomization (MR) analysis on the traits selected by TraitScan using the TwoSampleMR package (Hemani et al., 2018, 2017). The instrumental variables were selected from either UK Biobank or EWS GWAS data and were clumped by *r*^2^ *<* 0.001 and p-value *<* 5 *×* 10^*−*5^. For the selected traits with more than one instrumental variable, inverse-variance weighted (IVW), Egger regression, weighted median, simple mode, and weighted mode methods were applied, while the Wald ratio method was applied for the selected traits with one instrumental variable. Full results are reported in Table S2. EWS was shown to be causal for lower Alkaline phosphatase levels, the trait selected to be associated with SNP rs10822056 in TraitScan but not PheWAS (MR IVW coefficient = *−*0.53 with p-value = 0.084 and MR weighted median coefficient = *−*1.10 with p-value = 1.31 *×* 10^*−*4^). For the casual direction where EWS was the exposure, no MR test reached statistical significance after accounting for multiple testing, suggesting no causal effect from EWS to the selected UK Biobank traits.

## 4 Simulation

Throughout the simulations, we used five metrics to assess the performance of each method, i.e., power, size, recall, precision, and Jaccard similarity. Let *p* be the total number of traits, *S*^*∗*^ be the subset of traits as chosen by a particular method and let *S*_0_ be the true subset of pleiotropic traits. The size was defined as *E*| *S*^*∗*^|. Then, we defined precision to be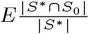, the proportion of traits identified by the method that was truly associated with the SNP. We defined recall to be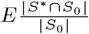, the proportion of the pleiotropic traits identified by the method. We defined Jaccard similarity, a combination of precision and recall, to be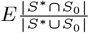. Finally, power was assessed by comparing the observed statistic to the simulated distribution of null statistics.

We compared the performance of variable selection and testing for TraitScan using summary statistics with some existing methods: PheWAS (with Bonferroni adjustment), CPASSOC (*S*_*hom*_, *S*_*het*_) (Zhu et al., 2015), MTaSPUs (Kim et al., 2015), and generalized higher criticism (GHC) (Barnett et al., 2017). Among these methods, MTaSPUs and *S*_*hom*_ cannot select traits, and thus we only evaluated their performance in terms of statistical power. *S*_*het*_ includes a parameter grid as the thresholds for z-scores and can naturally select out the traits with absolute z-scores smaller than each threshold. As suggested by their package, the parameter grid was set as the observed trait p-values. The GHC was originally proposed for SNP set association with a single trait, and here we used it for a single SNP association with multiple traits. We did not compare our method with ASSET because it is not computationally efficient in our simulation settings (706 traits for the real data variance-covariance scenario and 50 traits for the rest of the scenarios).

We conducted simulations under multiple scenarios to show TraitScan has higher power and Jaccard similarity under moderately sparse (|*S*_0_|*/p <* 0.4) and sparse situations (|*S*_0_|*/p <* 0.05). We focus the discussion on the scenarios with varying numbers of truly associated traits (scenario 1) or with real data variance-covariance matrix (scenario 2) and briefly discuss the other four scenarios and their results.

We assessed the method performance by varying the number of truly associated traits |*S*_0_| in scenario 1 (Figure 1). In terms of statistical power, we observed that TraitScan test statistics (HC, TC, HC+TC) had the highest power under moderately sparse and sparse situations (|*S*_0_| *<* 22, or |*S*_0_|*/p <* 0.44). The HC test was more powerful than the TC test under extremely sparse situations (|*S*_0_| = 1). When |*S*_0_| = 50, the GHC, MTaSPUs, PheWAS, and *S*_*hom*_ had the highest power, and TraitScan and *S*_*het*_ were less powerful. In terms of variable selection performance, we found that TraitScan test statistics (HC, TC, HC+TC) also had the highest Jaccard similarity under moderately sparse and sparse situations (|*S*_0_| *<* 22, or |*S*_0_|*/p <* 0.44), while *S*_*het*_ had betterJaccard similarity as increasing proportion of truly associated traits. However, *S*_*het*_ tended to over-select traits and thus had the lowest precision under most situations.

**Fig. 1:**
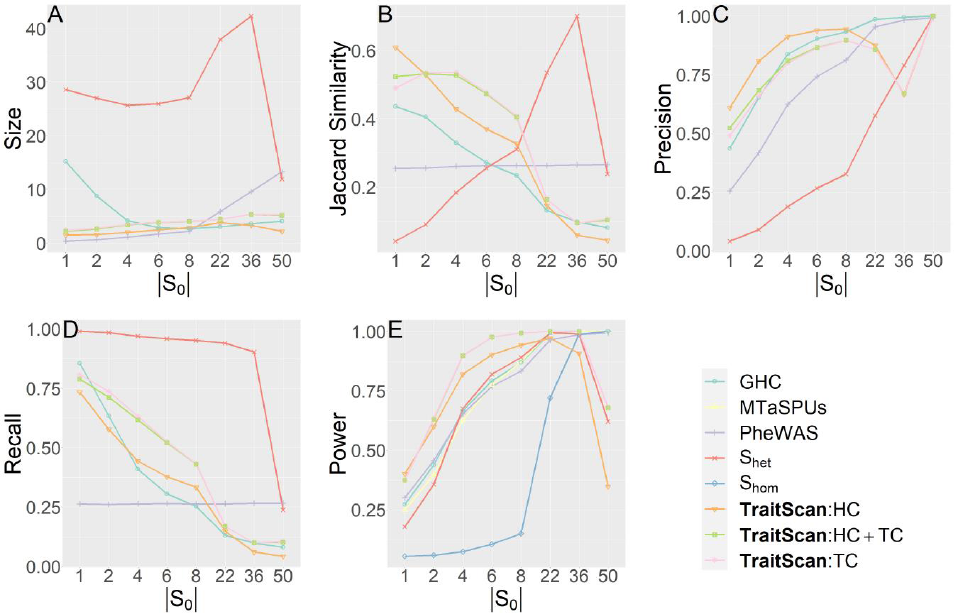
Simulation scenario 1 with varying numbers of truly associated traits (*p* = 50)

We also tested method performance using the covariance matrix and effect sizes estimated from the 706 UK biobank traits (Figure 2), where 38 traits with marginal p-value *≤* 0.05 were set as a true subset of pleiotropic traits. In this scenario, we compared the performance of PheWAS and TraitScan under trait-SNP associations of different strengths. *S*_*het*_, GHC, and MTaSPUs were not applied due to the computational burden, and *S*_*hom*_ was excluded due to the homogeneous effect assumption was not met in the simulation and unlikely to be met in real data analysis. By comparing the Type I errors (***β*** = **0**), we showed that TraitScan tests were well calibrated, while PheWAS had a slightly deflated Type I error rate due to its independence assumption among traits. Similar to scenario 1 mentioned above, TraitScan had higher power and Jaccard similarity over PheWAS under all effect sizes. We also notice that the selected trait size did not grow with effect size. When the effect sizes were small, the p-values of truly associated traits and null traits were close, and TraitScan tended to select a large set of traits as the most anonymous trait subset.

**Fig. 2:**
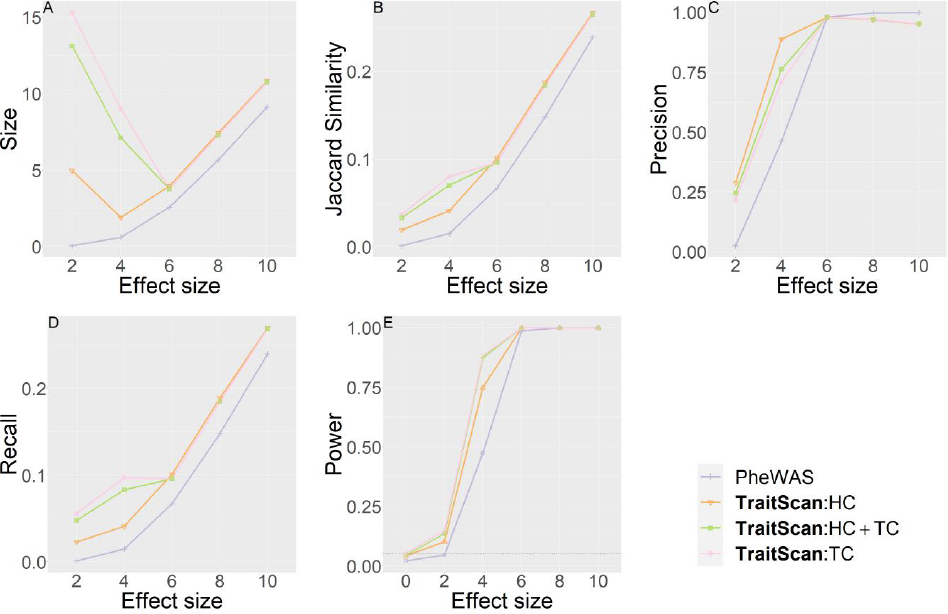
Simulation scenario 2 with real data covariance matrix and effects from SNP rs113663169 (*p* = 706, |*S*_0_| = 38). Effect size is in proportion to the estimated correlation observed in UK Biobank.

We showed that TraitScan can handle mixed types of traits (i.e., continuous and binary) simultaneously (Figure S1) and the performance followed the same pattern as continuous traits. TraitScan still demonstrated the highest power and Jaccard Similarity under effects varying in both directions and magnitudes (Table S3), under block-diagonal correlated traits (Figure S2), or different correlation magnitudes or structures (Table S4). Moreover, we found that TraitScan had higher power and Jaccard similarity when handling more highly correlated traits. The detailed parameter settings of the simulation can be found in Supplementary Materials.

## 5 Discussion

We proposed a new method called TraitScan for post-GWAS trait subset scanning and testing. While most of the existing multi-trait methods rely on domain knowledge, our method allows agnostic search among a large number of traits and is able to identify a set of traits with the most anomalousness. TraitScan utilizes the fast subset scan framework (Neill, 2012), resulting in a linear scan time over the number of traits. Taking correlation among traits into consideration, TraitScan has demonstrated higher power and trait selectivity than PheWAS when sparsity was low or modest. The method is compatible with both individual-level and summary-level GWAS data, although we focus more on summary-level GWAS data herein to allow an easy application to existing deeply phenotyped GWAS summary statistics databases.

In implementation, we recommend a grid of 200 *α*^*′*^*s* with the minimum *α* as the Bonferroni significant p-value cutoff and maximum *α* of 0.05. A practical issue faced by TraitScan and other threshold-based multi-trait methods is the choice of the density of thresholds. For the statistic *S*_*het*_, Zhu et al. (2015) recommended flexible thresholds which are the same as the input z-scores, while Bu et al. (2020) and TraitScan used fixed p-value thresholds. In simulations (not shown), we observed a dramatic power and precision loss of *S*_*het*_ using the same fixed threshold grid as in TraitScan. It is noteworthy that TraitScan always selects traits that are statistically significant in the decorrelated univariate analysis as we set the minimum *α* as the Bonferroni significant p-value cutoff. Based on our experience with simulations and real data analyses, we recommend a maximum *α* of 0.05 as traits with decorrelated p-values larger than 0.05 have never been selected by our algorithm. Since TraitScan includes a MC step simulating the null distribution of test statistics, an overly dense *α* grid may slow down the algorithm. We found a grid of 200 *α*^*′*^*s* is sufficiently dense when handling hundreds of GWAS traits.

Given a fixed number of *α*^*′*^*s*, TraitScan has an *O*(*p*) time complexity. If the number of *α*^*′*^*s* is proportional to *p*, the time complexity is *O*(*p*^2^). While for the other multi-trait analysis methods, the *S*_*het*_ test in CPASSOC also has an *O*(*p*) time complexity given a fixed p-value threshold, since it ranks the p-values and directly applies the threshold onto the raw p-values. When the thresholds are set as the input p-values, which is recommended in the CPASSOC pipeline, its time complexity is also *O*(*p*^2^). The ASSET has an *O*(2^*p*^) time complexity, meaning the computational time will be doubled once adding one more trait. Table 2 lists the computational time for analyzing 10 traits 5,000 times using different methods, including TraitScan with 10,000 iterations in the MC simulation (TraitScan-MC), TraitScan:HC test based on analytical null distribution (TraitScan-analytic), and TraitScan test using a precalculated null distribution estimated by MC with a given *p* (TraitScan-precalculated). The trait correlations and *β* parameters are the same as in Scenario 1.

**Table 2.**
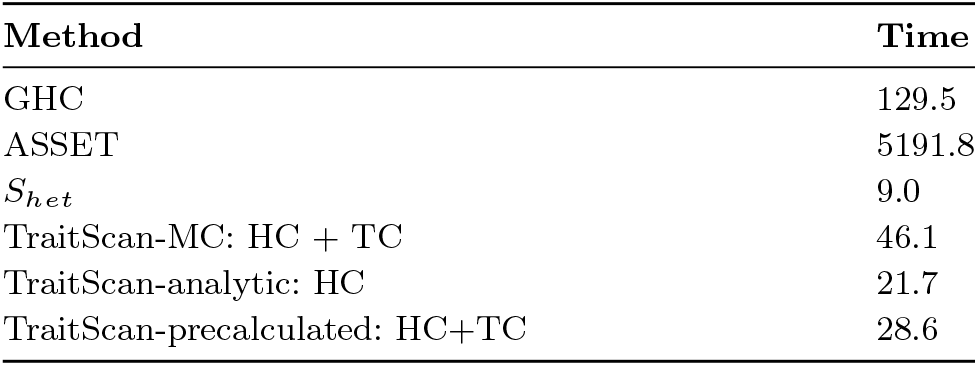
Computational time in seconds for 5,000 iterations (10 traits)

| Method | Time |
| --- | --- |
| GHC | 129.5 |
| ASSET | 5191.8 |
| $S_{het}$ | 9.0 |
| TraitScan-MC: HC + TC | 46.1 |
| TraitScan-analytic: HC | 21.7 |
| TraitScan-precalculated: HC+TC | 28.6 |

In the examination of traits associated with EWS, TraitScan identified eight additional trait-SNP associations which did not reach the PheWAS significance level. One of these traits, alkaline phosphatase, measured by blood assays, also showed significance in MR analysis, suggesting it was causally related to EWS. Evidence has shown the presence of abundant alkaline phosphatase activity in EWS tumor cells (Sharada et al., 2006); however, the direction of association between alkaline phosphatase and EWS was previously unknown. Another trait, IGF-1, was selected by TraitScan for SNPs rs2412476 on chromosome 15, rs6047482 on chromosome 20, and rs10822056 on chromosome 10. IGF1-receptor is known to be upregulated in EWS, and anti-IGF1 is an experimental therapy (Gonzalez et al., 2020). Besides, the SNP rs7742053, the only SNP that failed to reach genome-wide significance in both TraitScan and PheWAS, has recently been reported to have a specific role in the increased binding of GGAA microsatellite alleles with the chromosomal translocation encoding chimeric transcription factors (Lee et al., 2023).

Examining the genetic score of *KIZ* and *RREB1* allowed us to investigate whether any trait was associated with EWS on the gene level. If a gene is associated with the same set of traits, then likely multiple SNPs in the gene will be associated with the traits, leading to higher power than the single SNP test. However, if the weight in the genetic score is not informative, such as imputing gene expression in a non-relevant tissue, or if multiple SNPs in the gene suggest a different association with the trait, we would have diminished power. We did not observe any traits significantly associated with the genetic scores (i.e., imputed gene expression) of *KIZ* and *RREB1*, potentially because blood may not be the most relevant tissue for EWS. We note that like other methods, our results relied on the quality of GWAS data. When handling real GWAS data, we applied a couple of filtering steps to exclude traits that are not heritable. However, after the filtering steps, there were still a few traits that lacked reasonable explanations of their genetic heritability such as the inpatient record format, or the potential mechanisms to be associated with EWS, such as fruit intake within the past 24 hours. We suspect it was due to the inadequate adjustment for confounding in the original GWAS analysis (Holmes et al., 2019). Without accessing the individual-level data, it is difficult to examine or correct the summary-level GWAS data, although there is some recent work performing quality control on GWAS errors using summary statistics and a reference panel (Chen et al., 2021; Darrous et al., 2021).

To use TraitScan in real data analysis, the following additional steps could help avoid potential power loss and increase the interpretability of the results. First of all, we suggest removing highly correlated traits from the pool of putative traits by examining the empirical trait correlation matrix. The LTSS property of TraitScan requires traits to be independent of each other. As shown in the simulation, our method had relatively low statistical power when the genetic variant had the effects and correlations of the same direction on most of the traits. We find that the decorrelation step on z-scores shifted the means of z-scores of the truly associated traits towards zero, resulting in a power loss. Therefore, removing such traits could potentially improve the statistical power. Future work may be focused on developing subset algorithms balancing the computational time and scan sensitivity. Secondly, we recommend checking the correlation between the decorrelated traits and raw traits. Due to the trait decorrelation in TraitScan, trait selection and testing are performed on the decorrelated z-scores, which are essentially linear combinations of raw z-scores. Although the ZCA-cor decorrelation method maximizes the average correlation between each dimension of the decorrelated and original data, the decorrelated traits might be considered to differ from the original traits. Therefore, this step could improve the interpretability of the findings. In our real data analysis, 99% of the 706 UK Biobank traits had an empirical correlation with the original trait greater than 0.7.

The understanding of rare diseases such as childhood cancer has long been limited. TraitScan is able to provide a list of possible traits associated with EWS through the disease-linked genetic variants. As association does not imply causality, further biological experiments or additional data analysis approaches such as MR are required to study whether a trait and target disease are causally linked and whether the trait is a risk factor or a consequence of the disease.

## Data Availability

The summary-level GWAS data for UK Biobank can be downloaded through instructions at https://pan.ukbb.broadinstitute.org/downloads. The algorithm for the proposed work is packaged in R, available at https://github.com/RuiCao34/TraitScan.

https://pan.ukbb.broadinstitute.org/downloads

## Data Availability Statement

The summary level GWAS data for UK Biobank can be downloaded through instructions at https://pan.ukbb.broadinstitute.org/downloads. The algorithm for the proposed work is packaged in R, available at https://github.com/RuiCao34/TraitScan.

## Acknowledgments

The authors thank Prof. Jim Hodges of the Division of Biostatistics, University of Minnesota, who helped with administrative matters early in this project. The authors also want to thank Dr. Aubrey K. Hubbard and Mitchell J. Machiela from the Division of Cancer Epidemiology and Genetics at National Cancer Institute for sharing the summary statistics of Ewing Sarcoma. This work is supported by the Children’s Cancer Research Fund and by the Minnesota Supercomputing Institute at the University of Minnesota. Dr. Yang would like to further acknowledge St Baldrick Career Award for their support.

