## Supplementary material for "Subset scanning for multi-trait analysis using GWAS summary statistics": Supplmental Materials

#### 1 Significance of HC

As $F_{HC,\alpha}(S)$ is an decreasing function of $|S|$, for $\alpha = \{\alpha_1, \alpha_2, ..., \alpha_m\}$,

$$\begin{aligned} p_{HC} &= Pr(\max_\alpha \max_S F_{HC,\alpha}(S) \geqslant h_{HC}|H_0) \\ &= Pr(\max_\alpha \sqrt{\frac{1-\alpha}{\alpha} N_\alpha} \geqslant h_{HC}|H_0) \\ &= 1 - Pr(\max_\alpha \sqrt{\frac{1-\alpha}{\alpha} N_\alpha} < h_{HC}|H_0) \\ &= 1 - Pr(\bigcap_i^m \sqrt{\frac{1-\alpha_i}{\alpha_i} N_{\alpha_i}} < h_{HC}|H_0) \end{aligned} \tag{1}$$

**Lemma 1.** *For* $0 < \alpha_{(1)} < \alpha_{(2)} < ... < \alpha_{(m)} < 1$ *and* $j = 1, 2, ..., p$, $p_j^* \overset{iid}{\sim} Uniform(0, 1)$ *under the* $H_0$. $N_{\alpha_{(i)}} = \sum_{j=1}^p I(p_j^* < \alpha_{(i)})$ *follows a Binomial(p,* $\alpha_{(i)}$*) distribution. Moreover,* $N_{\alpha_{(i)}}|N_{\alpha_{(i+1)}} \sim Binomial(N_{\alpha_{(i+1)}}, \frac{\alpha_{(i)}}{\alpha_{(i+1)}})$*, which is independent of* $N_{\alpha_{(i+2)}}$, $N_{\alpha_{(i+3)}}$,...,$N_{\alpha_{(m)}}$.

Under the null,

$$Pr(\bigcap_{i}^{m}\sqrt{\frac{1-\alpha_i}{\alpha_i}N_{\alpha_i}} < h_{HC}) = Pr(\sqrt{\frac{1-\alpha_{(m)}}{\alpha_{(m)}}N_{\alpha_{(m)}}} < h_{HC})$$

$$Pr(\sqrt{\frac{1-\alpha_{(m-1)}}{\alpha_{(m-1)}}N_{\alpha_{(m-1)}}} < h_{HC}|\sqrt{\frac{1-\alpha_{(m)}}{\alpha_{(m)}}N_{\alpha_{(m)}}} < h_{HC})$$

$$Pr(\sqrt{\frac{1-\alpha_{(m-2)}}{\alpha_{(m-2)}}N_{\alpha_{(m-2)}}} < h_{HC}|\bigcap_{i}^{m}\sqrt{\frac{1-\alpha_{(i)}}{\alpha_{(i)}}N_{\alpha_{(i)}}} < h_{HC})$$

...

$$Pr(\sqrt{\frac{1-\alpha_{(1)}}{\alpha_{(1)}}N_{\alpha_{(1)}}} < h_{HC}|\bigcap_{i}^{m}\sqrt{\frac{1-\alpha_{(i)}}{\alpha_{(i)}}N_{\alpha_{(i)}}} < h_{HC})$$

We proposed the following algorithm to estimate $p_{HC}$:

**Step 1:** Calculate the marginal probabilities $Pr(N_{\alpha_{(m)}} = 1)$, ... ,$Pr(N_{\alpha_{(m)}} = \lceil \frac{\alpha_{(m)}}{1-\alpha_{(m)}} H_{HC}^2 - 1\rceil)$

**Step 2:** When $i = m-1$, calculate probabilities

$$Pr(N_{\alpha_{(i)}} = j|N_{\alpha_{(i+1)}} < \frac{\alpha_{(i+1)}}{1-\alpha_{(i+1)}}h_{HC}^2)$$

$$= \frac{Pr(\{N_{\alpha_{(i)}} = j\}\bigcap\{N_{\alpha_{(i+1)}} < \frac{\alpha_{(i+1)}}{1-\alpha_{(i+1)}}h_{HC}^2\})}{Pr(N_{\alpha_{(i+1)}} < \frac{\alpha_{(i+1)}}{1-\alpha_{(i+1)}}h_{HC}^2)}$$

$$= \frac{\sum_{k=j}^{\lceil \frac{\alpha_{(i+1)}}{1-\alpha_{(i+1)}}h_{HC}^2-1\rceil} Pr(\{N_{\alpha_{(i)}} = j\}\bigcap\{N_{\alpha_{(i+1)}} = k\})}{\sum_{k=j}^{\lceil \frac{\alpha_{(i+1)}}{1-\alpha_{(i+1)}}h_{HC}^2-1\rceil} Pr(N_{\alpha_{(i+1)}} = k)}$$

for each $0 \leq j < \frac{\alpha_{(i)}}{1-\alpha_{(i)}}h_{HC}^2$.

**Step 3:** When $i = m-2, ..., 1$, substitute probabilities of $N_{\alpha_{(i+1)}}$ by conditional probabilities, and $Pr(N_{\alpha_{(i)}} = j|\bigcup_{l=i+1}^{m}\{N_{\alpha_{(l)}} < \frac{\alpha_{(l)}}{1-\alpha_{(l)}}h_{HC}^2\})$ can be derived in sequential order by using the following method:

$$Pr(N_{\alpha_{(i)}} = j | \bigcup_{l=i+1}^{m} \{N_{\alpha_{(l)}} < \frac{\alpha_{(l)}}{1-\alpha_{(l)}} h^2_{HC}\})$$

$$= \frac{Pr(\{N_{\alpha_{(i)}} = j\} \bigcup \{N_{\alpha_{(i+1)}} < \frac{\alpha_{(i+1)}}{1-\alpha_{(i+1)}} h^2_{HC}\} | \bigcup_{l=i+2}^{m} \{N_{\alpha_{(l)}} < \frac{\alpha_{(l)}}{1-\alpha_{(l)}} h^2_{HC}\})}{Pr(N_{\alpha_{(i+1)}} < \frac{\alpha_{(i+1)}}{1-\alpha_{(i+1)}} h^2_{HC}) | \bigcup_{l=i+2}^{m} \{N_{\alpha_{(l)}} < \frac{\alpha_{(l)}}{1-\alpha_{(l)}} h^2_{HC}\}}$$

$$= \frac{\sum_{k=j}^{\lceil \frac{\alpha_{(i+1)}}{1-\alpha_{(i+1)}} h^2_{HC} - 1 \rceil} Pr(\{N_{\alpha_{(i)}} = j\} \bigcup \{N_{\alpha_{(i+1)}} = k\} | \bigcup_{l=i+2}^{m} \{N_{\alpha_{(l)}} < \frac{\alpha_{(l)}}{1-\alpha_{(l)}} h^2_{HC}\})}{\sum_{k=j}^{\lceil \frac{\alpha_{(i+1)}}{1-\alpha_{(i+1)}} h^2_{HC} - 1 \rceil} Pr(N_{\alpha_{(i+1)}} = k | \bigcup_{l=i+2}^{m} \{N_{\alpha_{(l)}} < \frac{\alpha_{(l)}}{1-\alpha_{(l)}} h^2_{HC}\})}$$

$$= \frac{\sum_{k=j}^{\lceil \frac{\alpha_{(i+1)}}{1-\alpha_{(i+1)}} h^2_{HC} - 1 \rceil} Pr(N_{\alpha_{(i)}} = j | N_{\alpha_{(i+1)}} = k, \bigcup_{l=i+2}^{m} \{N_{\alpha_{(l)}} < \frac{\alpha_{(l)}}{1-\alpha_{(l)}} h^2_{HC}\}) \quad Pr(N_{\alpha_{(i+1)}} = k | \bigcup_{l=i+2}^{m} \{N_{\alpha_{(l)}} < \frac{\alpha_{(l)}}{1-\alpha_{(l)}} h^2_{HC}\})}{\sum_{k=j}^{\lceil \frac{\alpha_{(i+1)}}{1-\alpha_{(i+1)}} h^2_{HC} - 1 \rceil} Pr(N_{\alpha_{(i+1)}} = k | \bigcup_{l=i+2}^{m} \{N_{\alpha_{(l)}} < \frac{\alpha_{(l)}}{1-\alpha_{(l)}} h^2_{HC}\})}$$

by Lemma 1,

$$= \frac{\sum_{k=j}^{\lceil \frac{\alpha_{(i+1)}}{1-\alpha_{(i+1)}} h^2_{HC} - 1 \rceil} Pr(N_{\alpha_{(i)}} = j | N_{\alpha_{(i+1)}} = k) Pr(N_{\alpha_{(i+1)}} = k | \bigcup_{l=i+2}^{m} \{N_{\alpha_{(l)}} < \frac{\alpha_{(l)}}{1-\alpha_{(l)}} h^2_{HC}\})}{\sum_{k=j}^{\lceil \frac{\alpha_{(i+1)}}{1-\alpha_{(i+1)}} h^2_{HC} - 1 \rceil} Pr(N_{\alpha_{(i+1)}} = k | \bigcup_{l=i+2}^{m} \{N_{\alpha_{(l)}} < \frac{\alpha_{(l)}}{1-\alpha_{(l)}} h^2_{HC}\})}$$

for each $0 \leq j < \frac{\alpha_{(i)}}{1-\alpha_{(i)}} h^2_{HC}$.

**Step 4:** For $i = m-1, ..., 1$

$$Pr(N_{\alpha_{(i)}} < \frac{\alpha_{(i)}}{1-\alpha_{(i)}} h^2_{HC} | N_{\alpha_{(i+1)}} < \frac{\alpha_{(i+1)}}{1-\alpha_{(i+1)}} h^2_{HC})$$

$$= \sum_{j=0}^{\lceil \frac{\alpha_{(i)}}{1-\alpha_{(i)}} h^2_{HC} - 1 \rceil} Pr(N_{\alpha_{(i)}} = j | N_{\alpha_{(i+1)}} < \frac{\alpha_{(i+1)}}{1-\alpha_{(i+1)}} h^2_{HC})$$

### 2 Real data analysis results

Table S1: EWS SNPs and UK Biobank traits

| SNP | Traits - Bonferroni | Traits - TraitScan (minp) | UK Biobank p-values |
|---|---|---|---|
| rs113663169 | Natural Hair color: blonde | Natural Hair color: blonde | $\leq 10^{-20}$ |
| | Natural Hair color: dark brown | Natural Hair color: dark brown | $4.44 \times 10^{-16}$ |
| | Natural Hair color: black | Natural Hair color: black | $2.34 \times 10^{-8}$ |
| | Comparative height size at age 10 | | $3.51 \times 10^{-7}$ |
| | Seated height | Seated height | $3.19 \times 10^{-7}$ |
| rs7742053 | | Glucose (blood) | $1.32 \times 10^{-2}$ |
| | | Fruit consumers (recent 24 hours) | $2.57 \times 10^{-2}$ |
| | | Alcohol usually taken with meals | $7.67 \times 10^{-3}$ |
| | | Breastfed as a baby | $1.74 \times 10^{-2}$ |
| | | Inguinal/femoral hernia repair | $1.25 \times 10^{-2}$ |
| | | Chest pain or discomfort | $1.90 \times 10^{-2}$ |
| | | Headaches for 3+ months | $2.05 \times 10^{-2}$ |
| | | Inpatient record: K63.4 * | $3.95 \times 10^{-2}$ |
| | | Main speciality of consultant: Urology | $1.97 \times 10^{-1}$ |
| | | Treatment speciality of consultant: Geriatric medicine | $9.77 \times 10^{-3}$ |
| | | Methods of admission to hospital: Transfer | $1.08 \times 10^{-1}$ |
| | | Sources of admission to hospital: Transfer to other NHS provider | $6.13 \times 10^{-2}$ |
| | | Inpatient record format: PEDW HES 2015 | $2.94 \times 10^{-2}$ |
| | | Non-accidental death in close genetic family | $3.21 \times 10^{-2}$ |
| | | Tea intake (cups per day) | $2.71 \times 10^{-2}$ |
| | | Duration walking for pleasure | $4.14 \times 10^{-2}$ |
| rs10822056 | Cystatin C (blood) | Cystatin C (blood) | $2.51 \times 10^{-9}$ |
| | Total protein (blood) | Total protein (blood) | $3.60 \times 10^{-7}$ |
| | Platelet crit (blood) | Platelet crit (blood) | $2.59 \times 10^{-13}$ |
| | Platelet distribution width (blood) | Platelet distribution width (blood) | $2.75 \times 10^{-8}$ |
| | Lymphocyte count (blood) | | $5.15 \times 10^{-9}$ |
| | Monocyte count (blood) | Monocyte count (blood) | $1.19 \times 10^{-13}$ |
| | Appendicectomy | Appendicectomy | $3.50 \times 10^{-6}$ |
| | Monocyte percentage (blood) | | $7.36 \times 10^{-6}$ |
| | Comparative height size at age 10 | | $9.23 \times 10^{-6}$ |
| | Ease of skin tanning | | $4.59 \times 10^{-6}$ |
| | Platelet count (blood) | | $1.06 \times 10^{-10}$ |
| | | Alkaline phosphatase (blood) | $5.62 \times 10^{-5}$ |
| rs2412476 | Albumin (blood) | Albumin (blood) | $3.93 \times 10^{-13}$ |
| | Alanine aminotransferase (blood) | Alanine aminotransferase (blood) | $1.33 \times 10^{-15}$ |
| | Aspartate aminotransferase (blood) | Aspartate aminotransferase (blood) | $\leq 10^{-20}$ |
| | Red blood cell (erythrocyte) count (blood) | Red blood cell (erythrocyte) count (blood) | $3.93 \times 10^{-10}$ |
| | Red blood cell (erythrocyte) distribution width (blood) | Red blood cell (erythrocyte) distribution width (blood) | $\leq 10^{-20}$ |
| | Mean corpuscular haemoglobin (blood) | | $2.37 \times 10^{-8}$ |
| | Gamma glutamyltransferase (blood) | | $1.16 \times 10^{-7}$ |
| | Reticulocyte percentage (blood) | | $1.18 \times 10^{-6}$ |
| | | Insulin-like growth factor 1 (blood) | $1.17 \times 10^{-4}$ |
| rs6047482 | Urea (blood) | Urea (blood) | $1.02 \times 10^{-7}$ |
| | | Insulin-like growth factor 1 (blood) | $1.68 \times 10^{-4}$ |
| rs10822056 | Insulin-like growth factor 1 | Insulin-like growth factor 1 (blood) | $6.16 \times 10^{7}$ |
| | | Knee pain experienced in last month | $7.57 \times 10^{-3}$ |
| | | Weight | $2.69 \times 10^{-4}$ |
| | | Red blood cell (erythrocyte) distribution width (blood) | $8.35 \times 10^{-4}$ |
| | | Monocyte percentage (blood) | $2.95 \times 10^{-4}$ |
| | | Seated height | $6.05 \times 10^{-6}$ |

* Diagnostic endoscopic examination of lower bowel and biopsy of lesion of lower bowel using fibreoptic sigmoidoscope.
** Coronary arteriography using two catheters.
*** Primary total prosthetic replacement of knee joint using cement.
**** Pain other than headache, facial pain, neck or shoulder pain,back pain, stomach or abdominal pain, hip pain, knee pain.

Table S2: Bidirectional MR analysis on traits selected by TraitScan

| UK Biobank Traits | Trait as exposure or outcome | Number of instru-mental SNPs | MR method | beta | se(beta) | p-value |
|---|---|---|---|---|---|---|
| Hair color (natural, before greying): Black | Exposure | 31 | MR Egger | -0.6253 | 1.5160 | 0.6831 |
| | | | Weighted median | -1.0850 | 1.3310 | 0.4149 |
| | | | Inverse variance weighted | -0.7064 | 1.2231 | 0.5636 |
| | | | Simple mode | 0.0317 | 3.6005 | 0.9930 |
| | | | Weighted mode | -0.4834 | 1.1944 | 0.6885 |
| | Outcome | 44 | MR Egger | 0.0018 | 0.0010 | 0.0846 |
| | | | Weighted median | -0.0004 | 0.0004 | 0.3037 |
| | | | Inverse variance weighted | 0.00003 | 0.0003 | 0.9135 |
| | | | Simple mode | -0.0014 | 0.0010 | 0.1469 |
| | | | Weighted mode | -0.0012 | 0.0008 | 0.1330 |
| Hair color (natural, before greying): Blonde | Exposure | 80 | MR Egger | -0.0022 | 0.7185 | 0.9976 |
| | | | Weighted median | 0.0403 | 0.7507 | 0.9572 |
| | | | Inverse variance weighted | -0.0704 | 0.5532 | 0.8987 |
| | | | Simple mode | 2.4800 | 1.9900 | 0.2164 |
| | | | Weighted mode | 0.0633 | 0.6689 | 0.9248 |
| | Outcome | 44 | MR Egger | -0.0034 | 0.0021 | 0.1056 |
| | | | Weighted median | -0.0005 | 0.0005 | 0.3438 |
| | | | Inverse variance weighted | -0.0009 | 0.0006 | 0.1535 |
| | | | Simple mode | -0.0006 | 0.0011 | 0.5864 |
| | | | Weighted mode | -0.0007 | 0.0011 | 0.5264 |
| Hair color (natural, before greying): Dark brown | Exposure | 77 | MR Egger | -0.2771 | 0.3567 | 0.4397 |
| | | | Weighted median | -0.0735 | 0.3781 | 0.8458 |
| | | | Inverse variance weighted | -0.0863 | 0.2978 | 0.7719 |
| | | | Simple mode | -1.1228 | 0.8654 | 0.1984 |
| | | | Weighted mode | -0.1664 | 0.3051 | 0.5870 |
| | Outcome | 44 | MR Egger | 0.0055 | 0.0028 | 0.0544 |
| | | | Weighted median | 0.0007 | 0.0008 | 0.4375 |
| | | | Inverse variance weighted | 0.0011 | 0.0008 | 0.1833 |
| | | | Simple mode | -0.0034 | 0.0024 | 0.1540 |
| | | | Weighted mode | 0.0028 | 0.0028 | 0.3133 |
| Sitting height | Exposure | 347 | MR Egger | -0.8993 | 0.5486 | 0.1021 |
| | | | Weighted median | -0.2166 | 0.3690 | 0.5572 |
| | | | Inverse variance weighted | 0.1853 | 0.2377 | 0.4355 |
| | | | Simple mode | 0.4762 | 0.8948 | 0.5949 |
| | | | Weighted mode | -0.2844 | 0.6382 | 0.6562 |
| | Outcome | 43 | MR Egger | -0.0041 | 0.0047 | 0.3892 |
| | | | Weighted median | -0.0013 | 0.0013 | 0.3204 |
| | | | Inverse variance weighted | 0.0010 | 0.0014 | 0.4502 |
| | | | Simple mode | -0.0014 | 0.0032 | 0.6795 |
| | | | Weighted mode | -0.0018 | 0.0036 | 0.6166 |
| Cystatin C (blood) | Exposure | 173 | MR Egger | 0.0516 | 0.2904 | 0.8593 |
| | | | Weighted median | 0.0587 | 0.2221 | 0.7916 |
| | | | Inverse variance weighted | 0.1226 | 0.2213 | 0.5794 |
| | | | Simple mode | 0.4161 | 0.9481 | 0.6613 |
| | | | Weighted mode | 0.1307 | 0.2113 | 0.5370 |
| | Outcome | 44 | MR Egger | -0.0020 | 0.0051 | 0.7044 |
| | | | Weighted median | 0.0004 | 0.0016 | 0.8180 |
| | | | Inverse variance weighted | -0.0003 | 0.0015 | 0.8271 |
| | | | Simple mode | 0.0013 | 0.0035 | 0.7200 |
| | | | Weighted mode | 0.0012 | 0.0029 | 0.6954 |
| Total protein (blood) | Exposure | 156 | MR Egger | 0.1103 | 0.6048 | 0.8556 |
| | | | Weighted median | 0.0737 | 0.4253 | 0.8625 |
| | | | Inverse variance weighted | -0.7298 | 0.2776 | 0.0086 |
| | | | Simple mode | -0.3546 | 0.9808 | 0.7181 |
| | | | Weighted mode | 0.5422 | 0.5112 | 0.2904 |
| | Outcome | 44 | MR Egger | -0.0010 | 0.0068 | 0.8872 |
| | | | Weighted median | 0.0000 | 0.0018 | 0.9927 |
| | | | Inverse variance weighted | 0.0003 | 0.0020 | 0.8994 |
| | | | Simple mode | -0.0005 | 0.0040 | 0.9026 |
| | | | Weighted mode | -0.0009 | 0.0040 | 0.8284 |
| Platelet crit (blood) | Exposure | 247 | MR Egger | 0.0029 | 0.3242 | 0.9928 |
| | | | Weighted median | 0.5875 | 0.2907 | 0.0433 |
| | | | Inverse variance weighted | 0.1649 | 0.1761 | 0.3491 |

| | | | | | | |
|---|---|---|---|---|---|---|
| | | | Simple mode | -0.0331 | 0.6970 | 0.9622 |
| | | | Weighted mode | 0.3769 | 0.3406 | 0.2695 |
| | Outcome | 44 | MR Egger | 0.0037 | 0.0052 | 0.4852 |
| | | | Weighted median | -0.0003 | 0.0017 | 0.8388 |
| | | | Inverse variance weighted | 0.0010 | 0.0015 | 0.4958 |
| | | | Simple mode | 0.0006 | 0.0033 | 0.8555 |
| | | | Weighted mode | 0.0015 | 0.0025 | 0.5566 |
| Platelet distribution width (blood) | Exposure | 188 | MR Egger | 0.3066 | 0.3345 | 0.3605 |
| | | | Weighted median | 0.1412 | 0.2883 | 0.6242 |
| | | | Inverse variance weighted | -0.0836 | 0.1960 | 0.6697 |
| | | | Simple mode | 0.2417 | 0.6272 | 0.7004 |
| | | | Weighted mode | 0.6521 | 0.3851 | 0.0920 |
| | Outcome | 44 | MR Egger | -0.0122 | 0.0070 | 0.0902 |
| | | | Weighted median | 0.0019 | 0.0017 | 0.2787 |
| | | | Inverse variance weighted | 0.0011 | 0.0022 | 0.6073 |
| | | | Simple mode | 0.0032 | 0.0038 | 0.3982 |
| | | | Weighted mode | 0.0030 | 0.0039 | 0.4410 |
| Monocyte count (blood) | Exposure | 189 | MR Egger | 0.1437 | 0.2995 | 0.6319 |
| | | | Weighted median | 0.2251 | 0.2702 | 0.4048 |
| | | | Inverse variance weighted | 0.2004 | 0.1711 | 0.2416 |
| | | | Simple mode | 0.4239 | 0.5588 | 0.4491 |
| | | | Weighted mode | 0.1787 | 0.2918 | 0.5411 |
| | Outcome | 44 | MR Egger | 0.0016 | 0.0062 | 0.7995 |
| | | | Weighted median | -0.0029 | 0.0018 | 0.1109 |
| | | | Inverse variance weighted | -0.0004 | 0.0018 | 0.8448 |
| | | | Simple mode | -0.0082 | 0.0037 | 0.0322 |
| | | | Weighted mode | -0.0063 | 0.0034 | 0.0690 |
| Appendicectomy | Exposure | 13 | MR Egger | -6.6517 | 31.5233 | 0.8367 |
| | | | Weighted median | -7.7238 | 5.4113 | 0.1535 |
| | | | Inverse variance weighted | 2.4130 | 8.8971 | 0.7862 |
| | | | Simple mode | -9.2114 | 8.1080 | 0.2781 |
| | | | Weighted mode | -8.5374 | 6.6316 | 0.2222 |
| | Outcome | 43 | MR Egger | -0.0006 | 0.0013 | 0.6457 |
| | | | Weighted median | 0.0001 | 0.0005 | 0.8164 |
| | | | Inverse variance weighted | 0.0003 | 0.0004 | 0.4689 |
| | | | Simple mode | 0.0004 | 0.0010 | 0.6513 |
| | | | Weighted mode | 0.0002 | 0.0008 | 0.7940 |
| Alkaline phosphatase (blood) | Exposure | 179 | MR Egger | -0.6701 | 0.3350 | 0.0470 |
| | | | Weighted median | -1.1006 | 0.2878 | 0.0001 |
| | | | Inverse variance weighted | -0.5320 | 0.2017 | 0.0084 |
| | | | Simple mode | 0.1317 | 0.6292 | 0.8345 |
| | | | Weighted mode | -0.7932 | 0.2666 | 0.0033 |
| | Outcome | 44 | MR Egger | -0.0052 | 0.0067 | 0.4413 |
| | | | Weighted median | -0.0001 | 0.0017 | 0.9606 |
| | | | Inverse variance weighted | 0.0026 | 0.0020 | 0.1988 |
| | | | Simple mode | 0.0005 | 0.0035 | 0.8803 |
| | | | Weighted mode | 0.0005 | 0.0032 | 0.8699 |
| Albumin (blood) | Exposure | 121 | MR Egger | -0.6388 | 0.6319 | 0.3141 |
| | | | Weighted median | -0.7600 | 0.4665 | 0.1032 |
| | | | Inverse variance weighted | -0.5293 | 0.2811 | 0.0597 |
| | | | Simple mode | -1.3933 | 1.1572 | 0.2310 |
| | | | Weighted mode | -1.8021 | 1.0915 | 0.1013 |
| | Outcome | 44 | MR Egger | 0.0018 | 0.0061 | 0.7750 |
| | | | Weighted median | -0.0010 | 0.0018 | 0.5797 |
| | | | Inverse variance weighted | -0.0006 | 0.0018 | 0.7508 |
| | | | Simple mode | -0.0036 | 0.0041 | 0.3887 |
| | | | Weighted mode | -0.0034 | 0.0038 | 0.3728 |
| Alanine aminotransferase (blood) | Exposure | 123 | MR Egger | 0.8396 | 0.6118 | 0.1725 |
| | | | Weighted median | 1.0285 | 0.4751 | 0.0304 |
| | | | Inverse variance weighted | 0.4225 | 0.2884 | 0.1429 |
| | | | Simple mode | -0.1479 | 1.0319 | 0.8862 |
| | | | Weighted mode | 1.0844 | 0.5640 | 0.0568 |
| | Outcome | 44 | MR Egger | -0.0092 | 0.0060 | 0.1348 |
| | | | Weighted median | -0.0009 | 0.0016 | 0.5541 |
| | | | Inverse variance weighted | -0.0026 | 0.0018 | 0.1449 |
| | | | Simple mode | -0.0013 | 0.0031 | 0.6789 |
| | | | Weighted mode | -0.0011 | 0.0025 | 0.6578 |
| Aspartate aminotransferase (blood) | Exposure | 141 | MR Egger | 0.9521 | 0.7622 | 0.2137 |
| | | | Weighted median | 0.8916 | 0.4574 | 0.0512 |

| | | | | | | |
|---|---|---|---|---|---|---|
| | | | Inverse variance weighted | 0.3029 | 0.3352 | 0.3662 |
| | | | Simple mode | -0.2635 | 1.0548 | 0.8031 |
| | | | Weighted mode | 1.0288 | 0.5381 | 0.0579 |
| | Outcome | 44 | MR Egger | -0.0055 | 0.0062 | 0.3812 |
| | | | Weighted median | -0.0015 | 0.0015 | 0.3449 |
| | | | Inverse variance weighted | -0.0013 | 0.0018 | 0.4798 |
| | | | Simple mode | -0.0020 | 0.0030 | 0.5156 |
| | | | Weighted mode | -0.0025 | 0.0027 | 0.3553 |
| Red blood cell (erythrocyte) count (blood) | Exposure | 191 | MR Egger | -0.0316 | 0.3999 | 0.9370 |
| | | | Weighted median | -0.2736 | 0.3970 | 0.4907 |
| | | | Inverse variance weighted | -0.3561 | 0.2213 | 0.1076 |
| | | | Simple mode | -1.2893 | 0.7981 | 0.1079 |
| | | | Weighted mode | -0.4695 | 0.3422 | 0.1718 |
| | Outcome | 44 | MR Egger | 0.0038 | 0.0045 | 0.3999 |
| | | | Weighted median | 0.0001 | 0.0015 | 0.9272 |
| | | | Inverse variance weighted | 0.0017 | 0.0013 | 0.1949 |
| | | | Simple mode | -0.0014 | 0.0035 | 0.6935 |
| | | | Weighted mode | -0.0009 | 0.0030 | 0.7648 |
| Red blood cell (erythrocyte) distribution width (blood) | Exposure | 162 | MR Egger | -0.0386 | 0.3596 | 0.9146 |
| | | | Weighted median | -0.1366 | 0.2854 | 0.6323 |
| | | | Inverse variance weighted | -0.1649 | 0.2057 | 0.4228 |
| | | | Simple mode | -0.8000 | 0.5948 | 0.1806 |
| | | | Weighted mode | -0.2294 | 0.3107 | 0.4613 |
| | Outcome | 44 | MR Egger | -0.0034 | 0.0072 | 0.6341 |
| | | | Weighted median | 0.0008 | 0.0018 | 0.6468 |
| | | | Inverse variance weighted | 0.0040 | 0.0021 | 0.0591 |
| | | | Simple mode | 0.0010 | 0.0032 | 0.7529 |
| | | | Weighted mode | 0.0006 | 0.0028 | 0.8296 |
| Insulin-like growth factor 1 (blood) | Exposure | 185 | MR Egger | 0.1786 | 0.4041 | 0.6589 |
| | | | Weighted median | 0.0229 | 0.3972 | 0.9540 |
| | | | Inverse variance weighted | -0.0933 | 0.2038 | 0.6471 |
| | | | Simple mode | 0.0536 | 0.8604 | 0.9504 |
| | | | Weighted mode | -0.0079 | 0.5082 | 0.9876 |
| | Outcome | 44 | MR Egger | -0.0027 | 0.0064 | 0.6754 |
| | | | Weighted median | -0.0015 | 0.0017 | 0.3694 |
| | | | Inverse variance weighted | -0.0007 | 0.0019 | 0.7257 |
| | | | Simple mode | -0.0002 | 0.0034 | 0.9416 |
| | | | Weighted mode | -0.0013 | 0.0027 | 0.6328 |
| Urea (blood) | Exposure | 103 | MR Egger | -0.2938 | 0.8311 | 0.7244 |
| | | | Weighted median | -0.8006 | 0.5002 | 0.1095 |
| | | | Inverse variance weighted | -0.5520 | 0.3433 | 0.1078 |
| | | | Simple mode | 0.6343 | 1.1136 | 0.5702 |
| | | | Weighted mode | 0.0244 | 0.7545 | 0.9743 |
| | Outcome | 44 | MR Egger | 0.0022 | 0.0045 | 0.6328 |
| | | | Weighted median | -0.0005 | 0.0016 | 0.7515 |
| | | | Inverse variance weighted | 0.0015 | 0.0013 | 0.2426 |
| | | | Simple mode | -0.0015 | 0.0032 | 0.6437 |
| | | | Weighted mode | -0.0011 | 0.0028 | 0.7100 |
| Pain type(s) experienced in last month: knee pain | Exposure | 9 | MR Egger | 7.4722 | 16.8820 | 0.6714 |
| | | | Weighted median | 3.7904 | 5.3789 | 0.4810 |
| | | | Inverse variance weighted | -0.1545 | 3.8506 | 0.9680 |
| | | | Simple mode | 8.2950 | 9.3377 | 0.4003 |
| | | | Weighted mode | 7.0612 | 8.9783 | 0.4543 |
| | Outcome | 43 | MR Egger | 0.0003 | 0.0016 | 0.8555 |
| | | | Weighted median | 0.0007 | 0.0006 | 0.2527 |
| | | | Inverse variance weighted | 0.0004 | 0.0005 | 0.4439 |
| | | | Simple mode | 0.0010 | 0.0011 | 0.3781 |
| | | | Weighted mode | 0.0010 | 0.0011 | 0.3700 |
| Weight | Exposure | 211 | MR Egger | 0.2253 | 0.8623 | 0.7941 |
| | | | Weighted median | 0.0580 | 0.4604 | 0.8998 |
| | | | Inverse variance weighted | 0.1580 | 0.3020 | 0.6009 |
| | | | Simple mode | -0.9420 | 1.2106 | 0.4374 |
| | | | Weighted mode | -0.0885 | 0.7575 | 0.9071 |
| | Outcome | 42 | MR Egger | -0.0054 | 0.0050 | 0.2892 |
| | | | Weighted median | 0.0005 | 0.0016 | 0.7591 |
| | | | Inverse variance weighted | 0.0015 | 0.0015 | 0.3379 |
| | | | Simple mode | 0.0009 | 0.0035 | 0.7991 |
| | | | Weighted mode | 0.0003 | 0.0035 | 0.9234 |
| Monocyte percentage (blood) | Exposure | 175 | MR Egger | -0.1763 | 0.3370 | 0.6015 |

| | | | | | | |
|---|---|---|---|---|---|---|
| | | | Weighted median | 0.0601 | 0.2985 | 0.8405 |
| | | | Inverse variance weighted | 0.1872 | 0.1875 | 0.3182 |
| | | | Simple mode | -0.0527 | 0.5423 | 0.9227 |
| | | | Weighted mode | 0.0082 | 0.2985 | 0.9780 |
| | Outcome | 44 | MR Egger | 0.0047 | 0.0044 | 0.2983 |
| | | | Weighted median | -0.0021 | 0.0016 | 0.1886 |
| | | | Inverse variance weighted | -0.0002 | 0.0013 | 0.8990 |
| | | | Simple mode | -0.0039 | 0.0030 | 0.1986 |
| | | | Weighted mode | -0.0039 | 0.0028 | 0.1673 |

### 3 Simulation Details and Results

#### 3.1 Simulation settings

The simulated data were generated as follows. The $n$ elements of the minor allele dose vector $x$ were generated from a binomial distribution with two trials and the probabilities equal to a certain minor allele frequency (MAF). Then the trait vector $y$ was generated from the multivariate normal distribution with mean vector $x\beta$ and covariance matrix $\mathbf{\Sigma}$. Binary traits were generated by determining whether $y_i$ is greater than a cutoff in simulations. We let the $\beta$ vector have $p$ elements with $d$ many non-zero elements. Larger $\beta$ element values correspond to stronger associations between the traits and the SNP. The null SNPs for trait correlation estimation were simulated in the same way except the effect sizes are all zero. For the correlation structure, we tested homogeneous correlation (scenarios 1 and 3), real data variance-covariance matrix using the 706 UK Biobank traits (scenario 2), non-homogeneous correlation (scenario 4), and block-diagonal correlation scenarios (scenarios 5 and 6). Under the homogeneous correlation scenarios, all non-diagonal entries of the $\mathbf{\Sigma}$ matrix were equal to $\rho$, i.e., all pairs of traits had the same correlation. Diagonal entries of $\mathbf{\Sigma}$ are set to one. Under non-homogeneous correlation scenarios, half traits were negatively correlated with the rest, that is, for $i \neq j$, $\mathbf{\Sigma}_{ij} = -\rho$ if $i + j$ was odd, and $\mathbf{\Sigma}_{ij} = \rho$ otherwise. Within-block correlations were homogeneous, and inter-block correlations were zero. Mimicking a typical GWAS, we fitted linear regression for continuous traits and logistic regression for binary outcomes for each SNP $x$ and obtained their summary statistics.

In scenarios 1 to 5, we set sample size $n = 1,000$, and SNP of interest has a MAF$= 0.2$, number of null SNPs $= 2,500$, cutoff of binary outcome equals 0.2, and multiple correlation structures between traits to test their effects on performance. In scenario 6, we also kept the sample size $n = 1,000$ and MAF$= 0.2$. Since 706 traits were evaluated in this setting, we increased the number of null SNPs to 141,200. We conducted 1,000 runs of the simulations at each setting for power, and let $B = 10,000$ for the MC simulation step.

The detailed simulation settings are described below:

- Scenario 1: a total of 50 continuous traits with homogeneous effect sizes and correlation were simulated. The number of truly associated traits $|S_0|$ was varied at 1/2/4/6/8/22/36/50 where the effect size of $S_0$, $\beta_t$, was kept at 0.15, and correlation between any pair of traits was set to 0.2, i.e., $\sigma_{ij} = 0.2$.

- Scenario 2: variance-covariance matrix and effect sizes mimicking the UK Biobank data were simulated. $S_0$ contained 38 traits with raw p-values smaller than 0.05. The true effects were simulated by the estimated correlations between the traits and SNP rs113663169 from UK Biobank multiplied by a factor of 0/2/4/6/8/10 to evaluate method performance under different strengths of associations.

- Scenario 3: simulation setting is the same as scenario 1 except that half of the traits are binary. Meanwhile, half of the truly associated traits were continuous, and the other half were binary.

- Scenario 4: non-homogeneous effect sizes on 25 continuous traits and 25 binary traits with $|S_0| = 4$ were simulated. Effect sizes of two truly associated traits (one continuous and one binary) $\beta_{t1} = 0.1$, while effect sizes of two others $\beta_{t2} = 0.2/0.3/0.4/-0.2/-0.3/-0.4$. $\sigma_{ij} = 0.2$ .

- Scenario 5: 50 continuous traits with $\beta_t = 0.1$ and a block-diagonal correlation matrix were simulated. The 50 traits were grouped into four blocks which separately contain 5, 10, 15, and 20 traits. The within-block correlation $\sigma_{ij} = 0.2$ for $i, j$ in the same block, and between-block correlation $\sigma_{ij} = 0$ for $i, j$ in different blocks.

- Scenario 6: varying correlation magnitudes and directions on 25 continuous traits and 25 binary traits were simulated. Effect sizes of four truly associated traits (two continuous and two binary) $\beta_t = 0.15$. For settings with homogeneous correlations, $\sigma_{ij} = 0.2/0.5/0.8$ for each $i, j$. For settings with non-homogeneous correlations, $\sigma_{ij} = 0.2/0.5/0.8$ if $i + j$ is even, and $\sigma_{ij} = -0.2/-0.5/-0.8$ if $i + j$ is odd.

#### 3.2 Supplementary simulation results

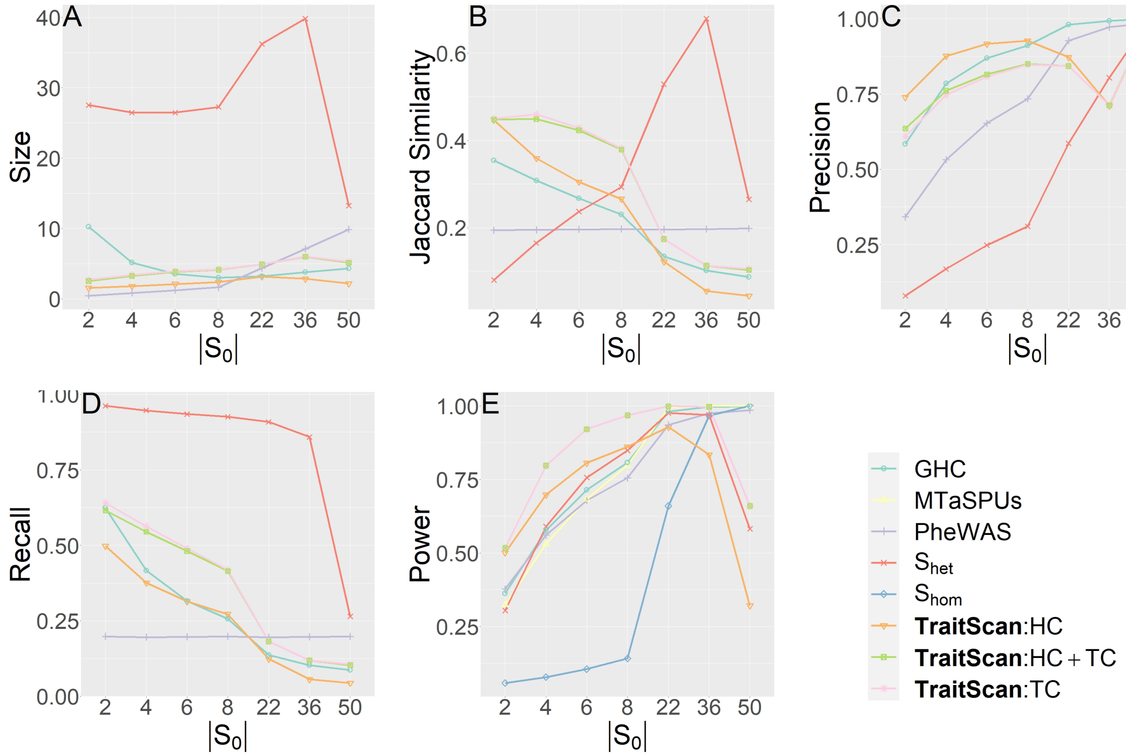


Figure S1: Simulations with continuous-binary mixed traits

Table S3: Simulation results for non-homogeneous effect sizes

| Non-homogeneous Effect Sizes | | 0.2 | 0.3 | 0.4 | -0.2 | -0.3 | -0.4 |
|---|---|---|---|---|---|---|---|
| Power | PheWAS | 0.741 | 0.996 | 1.000 | 0.755 | 0.994 | 1.000 |
| | $S_{hom}$ | 0.078 | 0.095 | 0.118 | 0.057 | 0.063 | 0.071 |
| | $S_{het}$ | 0.607 | 0.874 | 0.996 | 0.688 | 0.926 | 0.997 |
| | MTaSPUs | 0.697 | 0.993 | 1.000 | 0.715 | 0.995 | 1.000 |
| | GHC | 0.736 | 0.996 | 1.000 | 0.760 | 0.997 | 1.000 |
| | TraitScan-HC | 0.852 | 0.999 | 1.000 | 0.880 | 0.999 | 1.000 |
| | TraitScan-TC | 0.893 | 1.000 | 1.000 | 0.920 | 1.000 | 1.000 |
| | TraitScan-minp:HC+TC | 0.893 | 1.000 | 1.000 | 0.920 | 1.000 | 1.000 |
| Size | PheWAS | 1.072 | 1.944 | 2.130 | 1.040 | 1.900 | 2.122 |
| | $S_{het}$ | 25.438 | 19.232 | 11.904 | 23.998 | 17.705 | 11.269 |
| | GHC | 3.219 | 1.133 | 1.046 | 2.770 | 1.128 | 1.036 |
| | TraitScan-HC | 1.691 | 2.129 | 2.213 | 1.721 | 2.176 | 2.306 |
| | TraitScan-TC | 2.877 | 2.161 | 2.155 | 2.860 | 2.194 | 2.235 |
| | TraitScan-minp:HC+TC | 2.801 | 2.159 | 2.155 | 2.797 | 2.194 | 2.235 |
| Jaccard Similarity | PheWAS | 0.255 | 0.471 | 0.517 | 0.247 | 0.46 | 0.515 |
| | $S_{het}$ | 0.161 | 0.219 | 0.299 | 0.171 | 0.251 | 0.360 |
| | GHC | 0.288 | 0.278 | 0.262 | 0.298 | 0.277 | 0.259 |
| | TraitScan-HC | 0.363 | 0.497 | 0.515 | 0.384 | 0.515 | 0.547 |
| | TraitScan-TC | 0.428 | 0.497 | 0.512 | 0.455 | 0.515 | 0.539 |
| | TraitScan-minp:HC+TC | 0.424 | 0.497 | 0.512 | 0.451 | 0.515 | 0.539 |
| Precision | PheWAS | 0.716 | 0.98 | 0.986 | 0.732 | 0.979 | 0.986 |
| | $S_{het}$ | 0.172 | 0.281 | 0.468 | 0.179 | 0.291 | 0.473 |
| | GHC | 0.876 | 0.997 | 1.000 | 0.901 | 0.997 | 1.000 |
| | TraitScan-HC | 0.920 | 0.966 | 0.965 | 0.939 | 0.973 | 0.974 |
| | TraitScan-TC | 0.797 | 0.961 | 0.976 | 0.825 | 0.97 | 0.983 |
| | TraitScan-minp:HC+TC | 0.805 | 0.961 | 0.976 | 0.831 | 0.970 | 0.983 |
| Recall | PheWAS | 0.257 | 0.475 | 0.522 | 0.249 | 0.464 | 0.520 |
| | $S_{het}$ | 0.892 | 0.818 | 0.688 | 0.910 | 0.873 | 0.808 |
| | GHC | 0.344 | 0.279 | 0.262 | 0.346 | 0.278 | 0.259 |
| | TraitScan-HC | 0.375 | 0.507 | 0.526 | 0.394 | 0.523 | 0.556 |
| | TraitScan-TC | 0.501 | 0.509 | 0.520 | 0.523 | 0.524 | 0.545 |
| | TraitScan-minp:HC+TC | 0.493 | 0.508 | 0.520 | 0.516 | 0.524 | 0.545 |

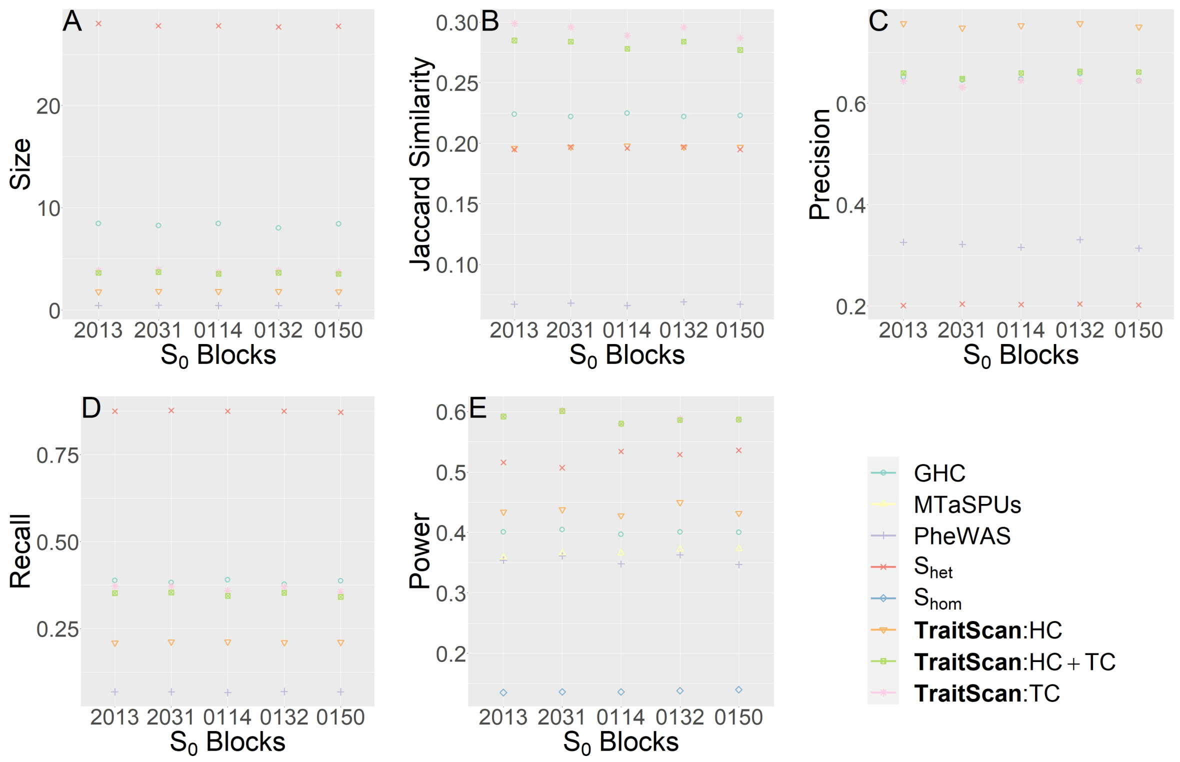


Figure S2: Simulations with block-correlated trait (x axis labels representing the numbers of truly associated traits in each block)

Table S4: Simulation results for different correlation magnitude and direction

| Non-homogeneous Correlation | | 0.2 | 0.3 | 0.4 | -0.2 | -0.3 | -0.4 |
|---|---|---|---|---|---|---|---|
| Power | PheWAS | 0.561 | 0.511 | 0.406 | 0.599 | 0.577 | 0.551 |
| | $S_{hom}$ | 0.078 | 0.059 | 0.060 | 0.317 | 0.430 | 0.740 |
| | $S_{het}$ | 0.590 | 0.656 | 0.681 | 0.651 | 0.730 | 0.869 |
| | MTaSPUs | 0.531 | 0.491 | 0.500 | 0.661 | 0.664 | 0.720 |
| | TraitScan-HC | 0.698 | 0.919 | 1.000 | 0.746 | 0.949 | 1.000 |
| | GHC | 0.577 | 0.429 | 0.266 | 0.613 | 0.479 | 0.326 |
| | TraitScan-TC | 0.797 | 0.960 | 1.000 | 0.831 | 0.974 | 1.000 |
| | TraitScan-minp:HC+TC | 0.797 | 0.960 | 1.000 | 0.831 | 0.974 | 1.000 |
| Size | PheWAS | 0.829 | 0.846 | 0.828 | 0.838 | 0.824 | 0.818 |
| | $S_{het}$ | 26.461 | 27.056 | 25.694 | 25.009 | 23.886 | 18.521 |
| | GHC | 5.144 | 10.563 | 22.127 | 4.464 | 7.153 | 14.026 |
| | TraitScan-HC | 1.802 | 2.117 | 3.044 | 1.833 | 2.245 | 3.046 |
| | TraitScan-TC | 3.407 | 3.178 | 2.945 | 3.366 | 3.048 | 2.950 |
| | TraitScan-minp:HC+TC | 3.261 | 3.117 | 2.945 | 3.234 | 3.002 | 2.950 |
| Jaccard Similarity | PheWAS | 0.532 | 0.487 | 0.395 | 0.573 | 0.559 | 0.541 |
| | $S_{het}$ | 0.169 | 0.208 | 0.280 | 0.175 | 0.230 | 0.399 |
| | GHC | 0.785 | 0.736 | 0.572 | 0.827 | 0.841 | 0.748 |
| | TraitScan-HC | 0.876 | 0.943 | 0.969 | 0.897 | 0.957 | 0.976 |
| | TraitScan-TC | 0.747 | 0.851 | 0.975 | 0.773 | 0.890 | 0.981 |
| | TraitScan-minp:HC+TC | 0.761 | 0.857 | 0.975 | 0.786 | 0.894 | 0.981 |
| Precision | PheWAS | 0.533 | 0.471 | 0.399 | 0.555 | 0.551 | 0.525 |
| | $S_{het}$ | 0.166 | 0.207 | 0.275 | 0.173 | 0.228 | 0.397 |
| | GHC | 0.785 | 0.731 | 0.577 | 0.817 | 0.838 | 0.733 |
| | TraitScan-HC | 0.875 | 0.944 | 0.965 | 0.895 | 0.958 | 0.975 |
| | TraitScan-TC | 0.704 | 0.841 | 0.953 | 0.743 | 0.880 | 0.966 |
| | TraitScan-minp:HC+TC | 0.742 | 0.869 | 0.965 | 0.775 | 0.902 | 0.974 |
| Recall | PheWAS | 0.196 | 0.201 | 0.199 | 0.198 | 0.196 | 0.196 |
| | $S_{het}$ | 0.947 | 0.939 | 0.904 | 0.95 | 0.949 | 0.932 |
| | GHC | 0.417 | 0.479 | 0.636 | 0.406 | 0.428 | 0.515 |
| | TraitScan-HC | 0.376 | 0.49 | 0.731 | 0.397 | 0.530 | 0.738 |
| | TraitScan-TC | 0.563 | 0.622 | 0.712 | 0.583 | 0.638 | 0.719 |
| | TraitScan-minp:HC+TC | 0.545 | 0.614 | 0.712 | 0.568 | 0.632 | 0.719 |
